# Clinical profile of scrub typhus patients during COVID pandemic: A sub-Himalayan single center experience

**DOI:** 10.1101/2022.08.20.22279010

**Authors:** Moirangthem Bikram Singh, Yogesh A. Bahurupi, Prasan Kumar Panda, Deepjyoti Kalita

**Author notes:** **Corresponding author** Prasan Kumar Panda, Asso. Professor, Dept. of Medicine, Sixth Floor, College Block, All India Institute of Medical Sciences (AIIMS), Rishikesh, India, 249203.

## Abstract

**Introduction:** Scrub typhus is tropical zoonotic disease, commonly presented with multi organ dysfunction and high mortality rate in untreated patients. This study was done to identify clinical features commonly associated with scrub typhus during COVID pandemics, parameters associated with severe scrub typhus and mortality.

**Methods:** This retrospective study was done in a tertiary care hospital with a total of 52 admitted scrub typhus positive patients in October 2020 to February 2022. Diagnosis was established by scrub IgM ELISA or Rapid antigen test. The clinical and laboratory data, duration of hospital stay and outcomes were collected. Common clinical and laboratory findings were of descriptive analysis. Factors associated with mortality were analysed using Chi-square test.

**Results:** Fever was the most common presenting symptoms on admission (94.2%) followed by respiratory abnormalities (38.46%). Acute kidney injury was the most common organ failure on admission (67.3%), followed by acute liver injury (46.2%) and thrombocytopenia (32.7%). MODS was seen in 46.15%. Of the total, 30.8% were admitted in ICU. Mortality was seen in 7.7% of all patients. On Chi-square analysis, altered mental status and coagulopathy were associated with significant mortality with p value <0.05.

**Conclusion:** Scrub typhus can manifest with potentially life-threatening complications such as acute kidney injury, acute liver injury, thrombocytopenia and MODS. The overall case-fatality rate was 7.7%, and presence of altered mental status and coagulopathy were associated with higher mortality. As per literature, COVID has changed few clinical profiles of scrub typhus compared to same center experience before.

## Introduction

Scrub typhus is a tropical infectious disease caused by *Orientia tsutsugamushi* which is a gram negative obligate intracellular organism whose polysaccharides bear an antigenic relationship to proteus OX-K that is thus used in serologic tests to confirm scrub typhus (1). The disease can result in severe arterial leakage and end-organ damage by causing disseminated vasculitis and perivascular inflammation and affect persons of all ages (2). It is quickly becoming a major public health problem as more cases are being detected in India. However, because it is both greatly underdiagnosed and underreported, its burden remains mysterious (3). It is endemic to India in Himalayan region and south and eastern part of India. most cases are reported during rainy season and post monsoon season (4). Diagnosis is often delayed due to overlapping clinical characteristics. Current diagnostic of choice is serology - scrub IgM ELISA in any patients presented with unspecific presentation of acute febrile illness particularly from endemic area.

Most common clinical manifestations are fever, headache, myalgia, body ache, nausea, vomiting, jaundice, chest pain, eschar, dry cough and shortness of breath (5)(6). Most common organ failures associated are acute kidney injury, acute liver injury, acute respiratory distress syndrome, acute meningoencephalitis, septic shock, myocarditis, unspecific arrythmias, thrombocytopenia and coagulopathy (7). Severities may vary from mild or asymptomatic patients to severe with multiorgan dysfunction. If the proper treatment is not given, the case fatality rate can range from 30 to 70 percent, while the median case fatality rate for treated patients is 1.4 percent and 6 percent, respectively (7).

This retrospective study aims to identify clinical features commonly associated with scrub typhus in this region, parameters associated with severe scrub typhus and mortality and ICU admission to aware clinicians about the severity of disease to facilitate in timely intervention and referral.

## Methodology

We conducted the study in a 1000 bedded hospital (tertiary level referral centre) in the north Indian state of Uttarakhand. The institutional ethics committee, AIIMS, Rishikesh, approved the study (AIIMS/IEC/20/720). The data collection was done through the e-medical records on the National Informatics Centre’s e-hospital portal used by the hospital and original file.

All adult scrub IgM ELISA and RAT positive patients admitted in AIIMS Rishikesh during the study period were included in the study. All other co-infections (like COVID, dengue, leptospirosis has been excluded from the study).

Data were collected regarding patient demographics, clinical features, vital parameters, laboratory data (complete blood counts, kidney function test, and liver function tests), chest radiography findings, duration of hospital stay, complications, and outcome on a predesigned data abstraction form. The nature and extent of organ dysfunction was also noted. Multiorgan dysfunction syndrome (MODS) was diagnosed as per established criteria MODS: simultaneous or sequential development of potentially reversible physiological derangement involving more than two organ systems (respiratory, cardiovascular, renal, hepatic, haematological, and neurological systems) (8).

The following outcomes were assessed:

1. Common clinical features on presentation and outcome

2. Organ failures on presentation

3. Parameters associated with mortality

The data were entered in the Microsoft excel spreadsheet, and analysis was done using IBM Statistical Package for Social Sciences (SPSS) version 25.0 (Chicago, US). Significance level were set at a level of p < 0.05 (two-tailed). Descriptive data with categorical variables were presented in number and percentage (%). Descriptive data with continuous variables were presented as mean ± standard deviation or median as per the normality of the data. For continuous variables, most data were converted to categorical variables for easier application form the published data as follows: TCL >12000/mm^3^, Hb<10mg/dl, platelet <100000mm^3^, urea >40mg/dl, creatinine >1.4mg/dl, hyponatremia Na <130meq/L, hyperkalaemia >5.2meq/L, transaminase > 3X ULN, Total bilirubin >2mg/dl, ALP >3XUNL and INR>1.2. Correlation between mortality outcome and baseline parameters was assessed by Chi-square test.

## Results

We identified 168 patients out of which 101 were excluded as other infective diagnosis was made and five were excluded because of exclusion. Remaining 52 patients were included in the study (Fig. 1).

**Fig 1:**
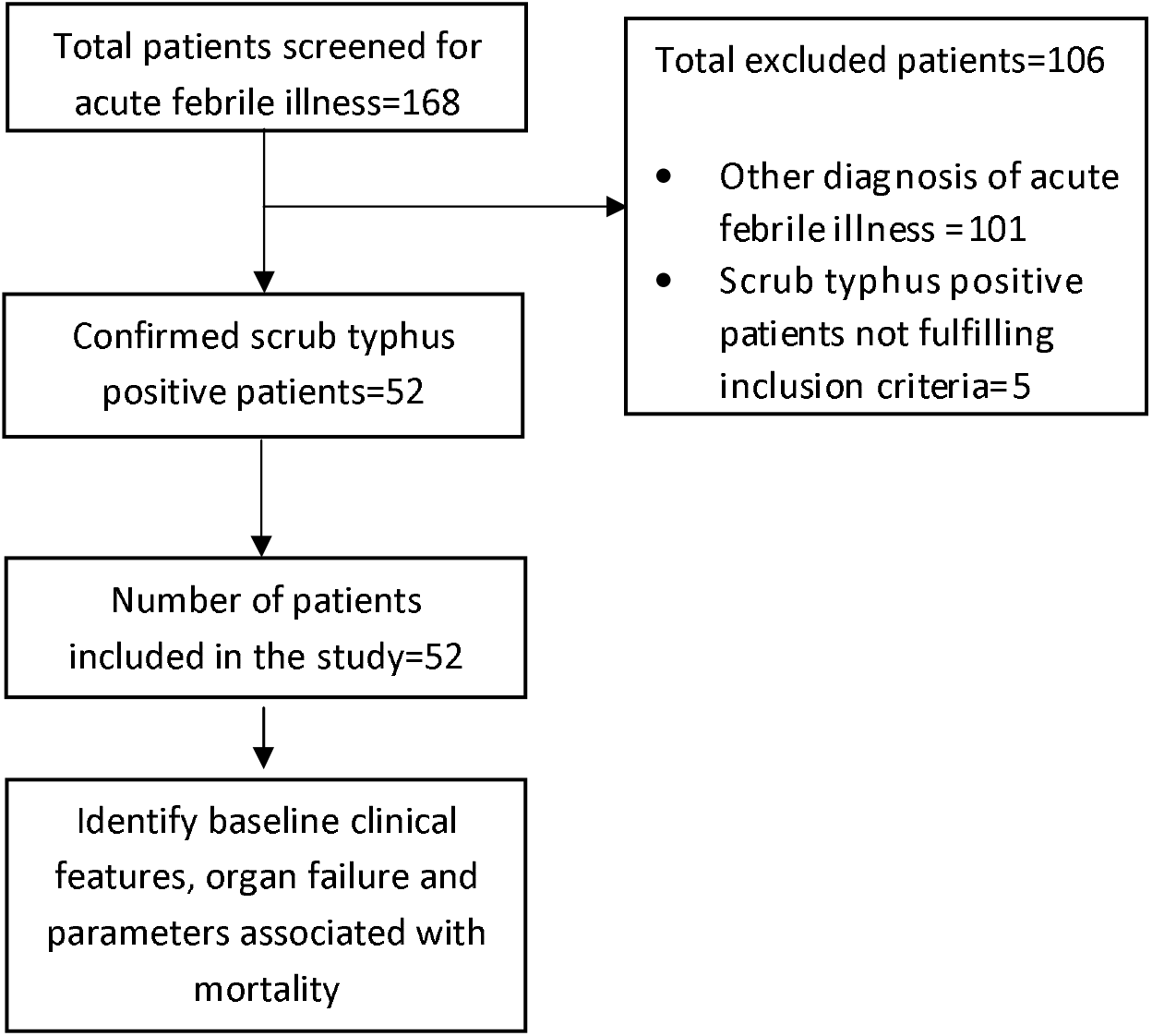
The study flow

Baseline characteristics of all the patients are shown in Table 1. Male predominance was seen. The mean age of the population is 46. Average length of hospital stay is one week. Average duration of fever before admission was 10. Occupational exposure and high risk behavior were found in very less number, may be due to COVID lockdown. In these 52 patients, 34 (65.4%) people had no co-morbidities; 14 had multiple co-morbidities and four had single co-morbidity. Most common comorbidity on admission was HTN (33.33%) followed by T2DM (22.2%). Most common pattern of fever on admission was intermittent. Majority were diagnosed by IgM ELISA with very less by unspecific RAT kit. Acute kidney injury was the most common organ failure on admission followed by acute liver injury and thrombocytopenia. MODS was seen in 24 (46.15%) of the total patients. One-third were admitted in ICU. Mortality was seen in few patients. Only two factors were associated with mortality: altered mental status (3 died out of 14 having) and coagulopathy (3 died out of 6 having) with p value <0.05.

**Table 1:**
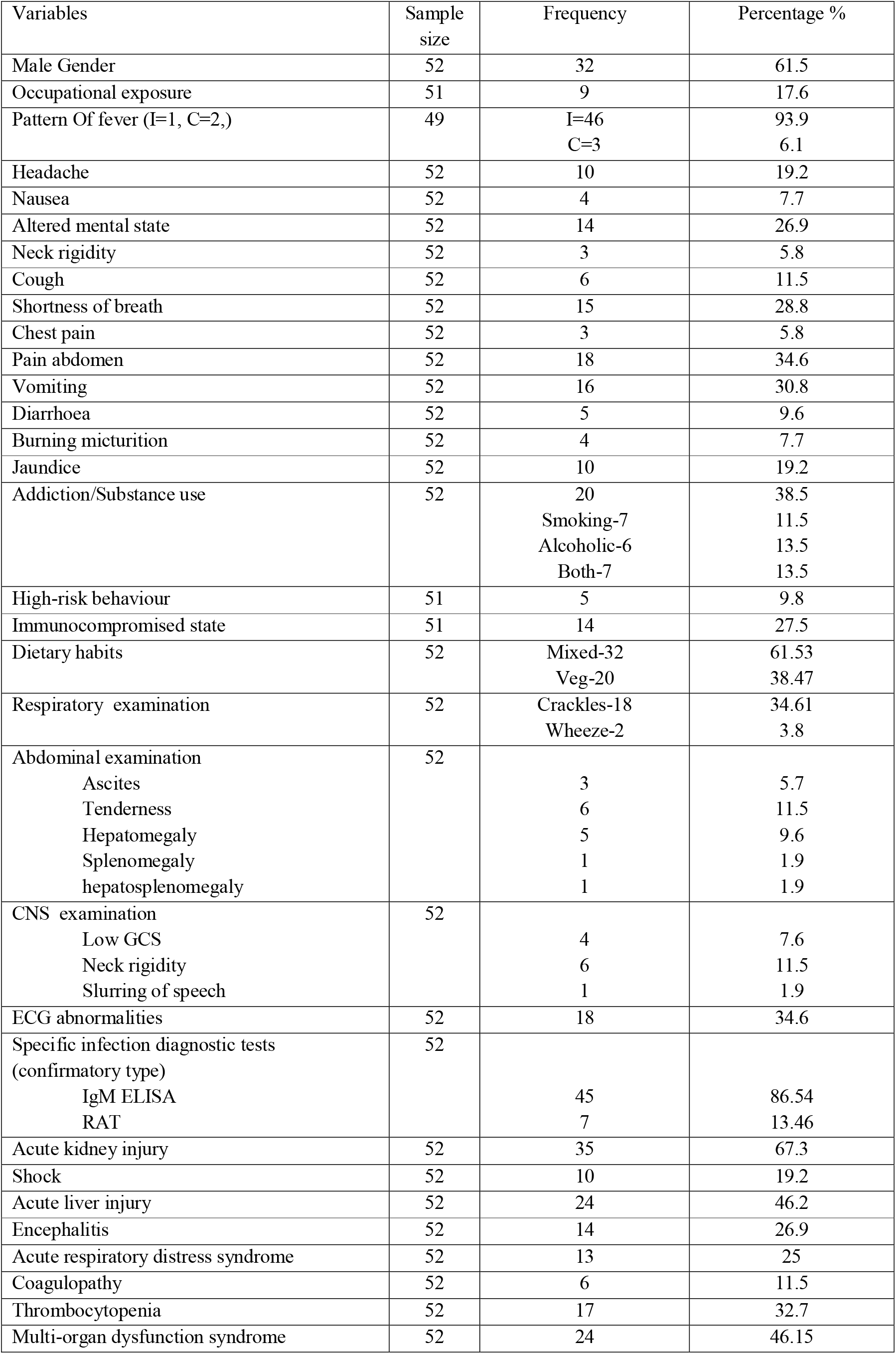
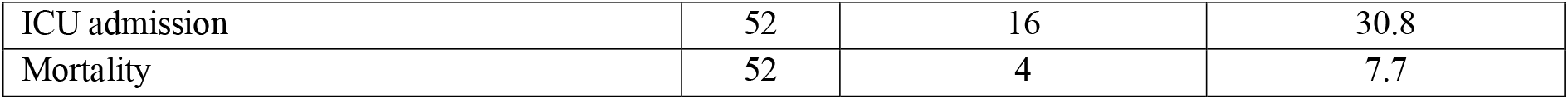
Baseline characteristics at admission of the participants.

| Variables | Sample size | Frequency | Percentage % |
| --- | --- | --- | --- |
| Male Gender | 52 | 32 | 61.5 |
| Occupational exposure | 51 | 9 | 17.6 |
| Pattern Of fever (I=1, C=2,) | 49 | I=46<br>C=3 | 93.9<br>6.1 |
| Headache | 52 | 10 | 19.2 |
| Nausea | 52 | 4 | 7.7 |
| Altered mental state | 52 | 14 | 26.9 |
| Neck rigidity | 52 | 3 | 5.8 |
| Cough | 52 | 6 | 11.5 |
| Shortness of breath | 52 | 15 | 28.8 |
| Chest pain | 52 | 3 | 5.8 |
| Pain abdomen | 52 | 18 | 34.6 |
| Vomiting | 52 | 16 | 30.8 |
| Diarrhoea | 52 | 5 | 9.6 |
| Burning micturition | 52 | 4 | 7.7 |
| Jaundice | 52 | 10 | 19.2 |
| Addiction/Substance use | 52 | 20<br>Smoking-7<br>Alcoholic-6<br>Both-7 | 38.5<br>11.5<br>13.5<br>13.5 |
| High-risk behaviour | 51 | 5 | 9.8 |
| Immunocompromised state | 51 | 14 | 27.5 |
| Dietary habits | 52 | Mixed-32<br>Veg-20 | 61.53<br>38.47 |
| Respiratory examination | 52 | Crackles-18<br>Wheeze-2 | 34.61<br>3.8 |
| Abdominal examination | 52 |  |  |
| Ascites |  | 3 | 5.7 |
| Tenderness |  | 6 | 11.5 |
| Hepatomegaly |  | 5 | 9.6 |
| Splenomegaly |  | 1 | 1.9 |
| hepatosplenomegaly |  | 1 | 1.9 |
| CNS examination | 52 |  |  |
| Low GCS |  | 4 | 7.6 |
| Neck rigidity |  | 6 | 11.5 |
| Slurring of speech |  | 1 | 1.9 |
| ECG abnormalities | 52 | 18 | 34.6 |
| Specific infection diagnostic tests<br>(confirmatory type) | 52 |  |  |
| IgM ELISA |  | 45 | 86.54 |
| RAT |  | 7 | 13.46 |
| Acute kidney injury | 52 | 35 | 67.3 |
| Shock | 52 | 10 | 19.2 |
| Acute liver injury | 52 | 24 | 46.2 |
| Encephalitis | 52 | 14 | 26.9 |
| Acute respiratory distress syndrome | 52 | 13 | 25 |
| Coagulopathy | 52 | 6 | 11.5 |
| Thrombocytopenia | 52 | 17 | 32.7 |
| Multi-organ dysfunction syndrome | 52 | 24 | 46.15 |
| ICU admission | 52 | 16 | 30.8 |
| Mortality | 52 | 4 | 7.7 |

**Table 2:** Baseline vitals and laboratory parameters at admission.

| **Table 2: Baseline vitals and laboratory parameters at admission** |  |  |  |  |  |
| --- | --- | --- | --- | --- | --- |
| Variables | N=52 | Mean | Std. Error of Mean | Median | SD |
| Age (in years) | 52 | 46.6 | 2.27 | 47 | 16.421 |
| Length of stay (days) | 52 | 7.87 | 0.53 | 7 | 3.85 |
| Days of fever | 51 | 9.47 | 0.80 | 8 | 5.728 |
| SBP (mmhg) | 52 | 109.67 | 2.92 | 110 | 21.115 |
| DBP (mmhg) | 52 | 71.08 | 1.83 | 72 | 13.235 |
| PR(/min) | 52 | 94.67 | 2.40 | 92.5 | 17.362 |
| RR (/min) | 52 | 21.06 | 0.51 | 20 | 3.707 |
| SPO2 (%) | 52 | 95.35 | 0.54 | 97 | 3.895 |
| Temperature chart (F) | 52 | 98.59 | 0.10 | 98.5 | 0.72208 |
| SGOT (U/L) | 51 | 208.21 | 26.56 | 180 | 189.736 |
| SGPT (U/L) | 51 | 144.73 | 18.21 | 105 | 130.045 |
| T. Bilirubin (mg/dl) | 52 | 2.73 | 0.41 | 1.33 | 3.024 |
| D. Bilirubin (mg/dl) | 52 | 1.75 | 0.29 | 0.82 | 2.154 |
| I. Bilirubin (mg/dl) | 52 | 0.97 | 0.13 | 0.675 | 0.985 |
| ALP (U/L) | 52 | 725.80 | 87.59 | 469.5 | 631.626 |
| GGT (U/L) | 52 | 177.52 | 23.83 | 107 | 171.841 |
| Albumin (g/dl) | 52 | 2.95 | 0.08 | 3.005 | 0.621 |
| Globulin (g/dl) | 52 | 2.85 | 0.10 | 2.73 | 0.749 |
| PT | 52 | 17.77 | 1.53 | 15.1 | 11.077 |
| INR | 52 | 1.342 | 0.11 | 1.15 | 0.838 |
| Urea(mg/dl) | 52 | 95.21 | 8.01 | 94.55 | 57.794 |
| creatinine(mg/dl) | 52 | 1.67 | 0.19 | 1.255 | 1.395 |
| Na+ (mmol/L) | 52 | 136.23 | 1.00 | 135 | 7.258 |
| K+(mmol/L) | 52 | 4.40 | 0.11 | 4.5 | 0.798 |
| Ca++ (mg/dl) | 52 | 8.01 | 0.20 | 7.9 | 1.478 |
| PO4- (mg/dl) | 52 | 3.20 | 0.16 | 2.95 | 1.212 |
| Uric acid (mg/dl) | 52 | 7.68 | 0.51 | 7.01 | 3.699 |
| Hb (g/dl) | 52 | 11.45 | 0.35 | 11.55 | 2.582 |
| TLC (/mm<sup>3</sup>) | 52 | 12.93 | 0.88 | 11.685 | 6.364 |
| PLT (/mm<sup>3</sup>) | 52 | 110.44 | 9.66 | 95 | 69.668 |

## Discussion

Scrub typhus is a potentially fatal disease. COVID has had marked impact in life style of the population, especially outdoor activities and hospitalisation for non-COVID illnesses. Considering these facts, this study has shown fever as the most common presentation in 93.9% which is similar to finding of Pathak et al (9)(10)(11). Mean age of the patients was 46.6 and average duration of hospital stay was 7.87 which is slightly lower than the finding of Varghese et al. (10). Eschar, which is considered as pathognomic of scrub typhus, wasn’t found in any of the patients, which is contrary to the most of the study finding showing 50-80% positivity in south Asian studies.

Most common organ failure on presentation was acute kidney injury in 67.3% but study finding of Pathak et al found that myocarditis was the most common 72.4% (9). This finding was also different from the finding of Griffith et al where respiratory (96.6%) and haematological (86.2%) dysfunction were most frequent (12). In the study of Jose A et al in the hospital of present study, thrombocytopenia was most common (56%) f/b liver injury (50%) compared to 32.7% and 46.2% respectively in this study (13). This may be effect of COVID pandemic. MODS was seen in 46.15% which is much lower than the study of Grifith et al having 85.2% but higher than another study by Bonnell A et al having 24.1% (12)(14).

Mortality rate of 7.7% was seen in present study which is close to the finding of Devasagayam et al in the systematic review of 18,781 patients with mortality rate of 7.3% (3). Among all the parameters analysed for association with mortality, only presence of altered mental status and coagulopathy were significantly associated with p value <0.05 which was different from the study finding of Adhikari et al that showed HR >100/min, SBP < 90mmHg, serum creatinine > 1.4mg/dl, kidney injury requiring dialysis, acute respiratory distress syndrome and shock requiring vasopressor leading higher mortality. In the study of Varghese et al, shock requiring vasoactive agents, CNS dysfunction, and renal failure was associated with mortality. These are also few observations that shows COVID has changed some patterns in clinical profile of scrub typhus.

The study has some limitations. It was conducted in a tertiary care hospital which is a referral centre, so present finding may not reflect the presentation at periphery health centre or community. The diagnosis was done using IgM ELISA and rapid antigen testing which are not gold standard diagnostic technique but clinical with supportive laboratory evidences confirmed it to be scrub typhus.

## Conclusion

During COVID era, scrub typhus presents mostly with fever (94.2%) followed by respiratory abnormalities (38.46%). At admission, most common organ failure was acute kidney injury (67.3%), followed by acute liver injury (46.2%) and thrombocytopenia (32.7%). MODS was seen in half of the patients while mortality was in 7.7%. Presence of altered mental status and coagulopathy (deranged INR) should prompt referral to higher centre as they are associated with mortality. By comparison with pre-COVID literatures, COVID has changed few clinical profiles.

## Data Availability

It will be made available to others as required upon requesting the corresponding author.

## Contributors

All authors contributed to the data collection, data analysis, and manuscript writing, critically reviewed the draft, and approved it for publication.

## Data sharing

It will be made available to others as required upon requesting the corresponding author.

## Acknowledgment

COVID care team was collecting data, special thanks to all of them.

## Conflicts of interest

We declare that we have no conflicts of interest.

## Funding source

None

## Notes

### Competing Interest Statement

The authors have declared no competing interest.

### Author Declarations

The institutional ethics committee, AIIMS, Rishikesh, approved the study (AIIMS/IEC/20/720).

## References

1. Rapsang AG, Bhattacharyya P. Scrub typhus. Indian J Anaesth [Internet]. 2013 Mar [cited 2022 Jul 11];57(2):127. Available from: /pmc/articles/PMC3696258/

2. Budha O, Singh C, Panda P. Scrub Typhus. 2020;(June).

3. Devasagayam E, Dayanand D, Kundu D, Kamath MS, Kirubakaran R, Varghese GM. The burden of scrub typhus in India: A systematic review. PLoS Negl Trop Dis [Internet]. 2021 Jul 1 [cited 2022 May 2];15(7):e0009619. Available from: https://journals.plos.org/plosntds/article?id=10.1371/journal.pntd.0009619

4. Lakshmi Rmmvn, Dharma TV, Sudhaharan S, Surya SMV, Emmadi R, Yadati SR, et al. Prevalence of scrub typhus in a tertiary care centre in Telangana, south India. Iran J Microbiol [Internet]. 2020 [cited 2022 May 2];12(3):204. Available from: /pmc/articles/PMC7340608/

5. Kim DM, Kim SW, Choi SH, Yun NR. Clinical and laboratory findings associated with severe scrub typhus. BMC Infect Dis. 2010;10:0–6.

6. Pathania M, Amisha, Malik P, Rathaur VK. Scrub typhus: Overview of demographic variables, clinical profile, and diagnostic issues in the sub-Himalayan region of India and its comparison to other Indian and Asian studies. J Fam Med Prim Care [Internet]. 2019 [cited 2022 May 13];8(3):1189. Available from: /pmc/articles/PMC6482722/

7. Xu G, Walker DH, Jupiter D, Melby PC, Arcari CM. A review of the global epidemiology of scrub typhus. PLoS Negl Trop Dis. 2017;

8. Varghese GM, Trowbridge P, Janardhanan J, Thomas K, Peter J V., Mathews P, et al. Clinical profile and improving mortality trend of scrub typhus in South India. Int J Infect Dis. 2014 Jun 1;23:39–43.

9. Pathak S, Chaudhary N, Dhakal P, Shakya D, Dhungel P, Neupane G, et al. Clinical profile, complications and outcome of scrub typhus in children: A hospital based observational study in central Nepal. PLoS One [Internet]. 2019 Aug 1 [cited 2022 Jul 12];14(8):e0220905. Available from: https://journals.plos.org/plosone/article?id=10.1371/journal.pone.0220905

10. Varghese GM, Trowbridge P, Janardhanan J, Thomas K, Peter J V., Mathews P, et al. Clinical profile and improving mortality trend of scrub typhus in South India. Int J Infect Dis [Internet]. 2014;23:39–43. Available from: http://dx.doi.org/10.1016/j.ijid.2014.02.009

11. Nanda S, Varma M, Vidyasagar S. Clinical profile of scrub typhus. Int J Infect Dis [Internet]. 2012 Jun 1 [cited 2022 Jul 12];16:e267. Available from: http://www.ijidonline.com/article/S1201971212010697/fulltext

12. Griffith M, Peter J, Karthik G, Ramakrishna K, Prakash JA, Kalki R, et al. Profile of organ dysfunction and predictors of mortality in severe scrub typhus infection requiring intensive care admission. Indian J Crit Care Med. 2014;18(8):497–502.

13. Jose A, Chaudhary A, Panda P, Kalita D. Scrub typhus in the Himalayan ranges and sub-Himalayan plains: Recognising an expanding clinical syndrome. J Med Evid. 2020;1(1):8.

14. Bonell A, Lubell Y, Newton PN, Crump JA, Paris DH. Estimating the burden of scrub typhus: A systematic review. PLoS Negl Trop Dis [Internet]. 2017 Sep 25 [cited 2022 Apr 24];11(9):e0005838. Available from: https://journals.plos.org/plosntds/article?id=10.1371/journal.pntd.0005838

